# Estimating the distribution of time to extinction of infectious diseases in mean-field approaches

**DOI:** 10.1101/2020.07.10.20150359

**Authors:** Maryam Aliee, Kat S. Rock, Matt J. Keeling

## Abstract

A key challenge for many infectious diseases is to predict the time to extinction under specific interventions. In general this question requires the use of stochastic models which recognise the inherent individual-based, chance-driven nature of the dynamics; yet stochastic models are inherently computationally expensive, especially when parameter uncertainty also needs to be incorporated. Deterministic models are often used for prediction as they are more tractable, however their inability to precisely reach zero infections makes forecasting extinction times problematic. Here, we study the extinction problem in deterministic models with the help of an effective “birth-death” description of infection and recovery processes. We present a practical method to estimate the distribution, and therefore robust means and prediction intervals, of extinction times by calculating their different moments within the birth-death framework. We show these predictions agree very well with the results of stochastic models by analysing the simplified SIS dynamics as well as studying an example of more complex and realistic dynamics accounting for the infection and control of African sleeping sickness (*Trypanosoma brucei gambiense*).

## 1 Introduction

For many infectious diseases the eventual aim of control measures is eradication completely removing the pathogen from host populations and the environment. This has been accomplished for just two infections – smallpox and rinderpest – but is the target for a number of other diseases including polio, Guinea Worm and Yaws [1–3]. A key question is, therefore, predicting the time to eradication, which has clear implications for the likely duration and hence cost of any control program.

Markov-chain models are often used to study the spread of infectious diseases in a population [4, 5]. In such approaches the whole population is partitioned into compartments and system dynamics are described by rates of exchange between compartments.

Both stochastic and deterministic versions of such models have been developed to study dynamics of a range of infectious diseases. The results of deterministic and stochastic versions of the same model generally agree well for large compartment populations, but start to deviate when there are either low numbers of infections, or for small population sizes [4, 6–9].

Deterministic models provide a mean-field approximation of infection dynamics described by ordinary differential equations (ODEs). The steady states and time evolution of these models are tractable and typically well-behaved in the whole phase space. Various analytical and numerical methods are available to solve a set of ODEs; this makes deterministic models computationally efficient and therefore allow the exploration of large regions of parameter space relatively quickly. This is particularly important for fitting such models to available data in order to estimate the underlying mechanistic parameters.

Deterministic descriptions are not very practical, however, when it comes to studying rare events or very small populations. Importantly, the endpoint of an infection (when infection is eradicated) is by definition ambiguous in these models since the populations are represented by continuous variables that never reach zero. In practice, a proxy threshold is needed to determine when extinction may occur, however, it is not clear how such a threshold should be defined. Infection dropping below one may seem a natural measure, but in the literature different threshold values are applied [10, 11] generating shifts in the predicted extinction times.

In such circumstances, stochastic models are more favourable. In these models, transitions between compartments are generated by stochastic events, whose average is well captured by the mean-field image, but express divergent behaviours individually. These events and accordingly system state are specified by probability functions allowing for uncommon incidents. Stochastic models are generally computationally expensive; however, they come with a good deal of flexibility and capabilities that we would like to take the advantage of to understand the extinction of diseases.

In this manuscript, we will develop a basic framework to study the peri-elimination phase for an infectious disease using a simplified SIS (Susceptible-Infected-Susceptible) model that takes into account susceptible and infected compartments. We use solutions of the corresponding forward Kolmogorov equation to predict the point of infection extinction in deterministic solutions. We then extend this framework to estimate the extinction time of more complicated infection dynamics. Whilst other studies have indeed addressed similar questions around computation of mean extinction times of epidemiological or other physical systems [12–20], the present study focuses on the computation of the distribution in predicted extinction times which has not been done elsewhere. This aspect is crucially important as the policy implications of extinction occurring within shorter or longer periods could lead to different decision making. Indeed uncertainty in model prediction is not accounted for often enough in epidemiological modelling studies [21]. Representation of uncertainty allows the modeller to answer policy-driven questions such as “at what time would we expect there to be a *>*90% likelihood that elimination would be met?” or “what is the earliest we could hope to achieve zero infections?”, which the mean value does not address.

For this study, we are motivated by a desire to analyse the endpoint of African sleeping sickness (*Gambiense* human African trypanosomiasis; gHAT), which is a vector-borne disease transmitted to humans by tsetse [22–25]. Humans transition through different stages of the disease and can either be diagnosed and recover (through treatment) or typically die without it. Medical intervention campaigns have had great success in bringing down the burden of this disease - from 37,385 in the most recent peak in 1998 to a record low of 953 cases in 2018 [26]. To continue the drive forwards, the World Health Organization (WHO) has a goal to achieve global elimination of transmission (EOT) of sleeping sickness by 2030 [27]. Quantitative methods provide a way to forecast the expected time to meet the goal under a variety of strategies, however there is a strong need for refinement of the methodology to analyse the distribution of extinction times in a more rigorous mathematical framework. Previous studies have used different threshold criteria for the endpoint for gHAT, by considering when the incidence of new infections falls below one of: 1 per 10,000, 1 per 100,000, less than 0.5 new infections across the entire health zone (*∼*100,000) per year, even to a more severe threshold of *<*1 per 1,000,000 [23, 28–30].

## 2 Model Structure

The simple SIS model is a one-dimensional characterisation of an infectious disease, in which recovery (or by treatment or otherwise) quickly leaves the individual susceptible to further infection [4, 5]. For a population of size *N*, the number of infected individuals can be expressed as:

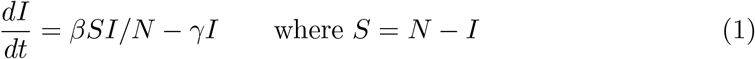

Here *β* is the transmission rate, *γ* is the recovery rate and *S* and *I* are the numbers of susceptible and infectious individuals respectively. Another important metric, *R*_0_ = *β/γ*, is the basic reproduction number. When *R*_0_ *>* 1 this model exhibits sigmoidal growth to the non-zero equilibrium (*I\** = 1 *−* 1*/R*_0_); whereas when *R*_0_ *<* 1 we observe exponential decay of infection to zero. Through changes to the transmission or recovery rates it is possible to modify the *effective reproduction number, R*_eff_ and push it below this critical threshold. It is this latter case that is of greatest interest here, as we are most concerned with diseases that are bound for eventual extinction (assuming controls remain in place and *R*_eff_ continues to be *<*1).

The non-linear *SI* term in equation (1) precludes many analytical results; however, if we are purely interested in extinction it is acceptable to assume that the number of infected individuals is relatively small and hence that *S ≈ N*. In this limit, we obtain a simple birth-death process for infections, where birth corresponds to infecting someone and death refers to recovery [31]

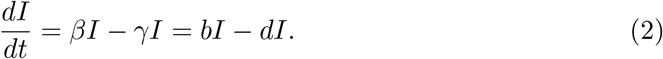

Here *β* plays the role of the *per capita* birth rate (*b*) and *γ* is equivalent to the *per capita* death rate (*d*). Throughout the rest of this paper we will use the parameters *b* and *d* to stress that we are approximating the dynamics as a birth-death process. Our condition that *R*_eff_ *<* 1 now corresponds to *d > b*.

### 2.1 Dynamics without births

We first consider the simplest case where *b* = *β* = 0, and so each infected individual recovers (or is treated) at a Poisson rate *d*, but no new infections occur. Starting with *n* infected individuals, the expected time for the first individual to recover is 1*/*(*dn*). After this recovery event, the population then has *n −* 1 infected individuals and the process can be iterated giving an expected time to extinction of:

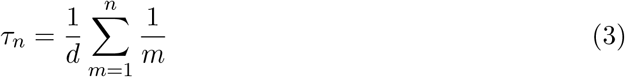

Figure 1A illustrates how the average extinction time *τ*_*n*_ depends on the initial number of infected people *n*, showing a logarithmic scaling for large *n*.

**Figure 1:**
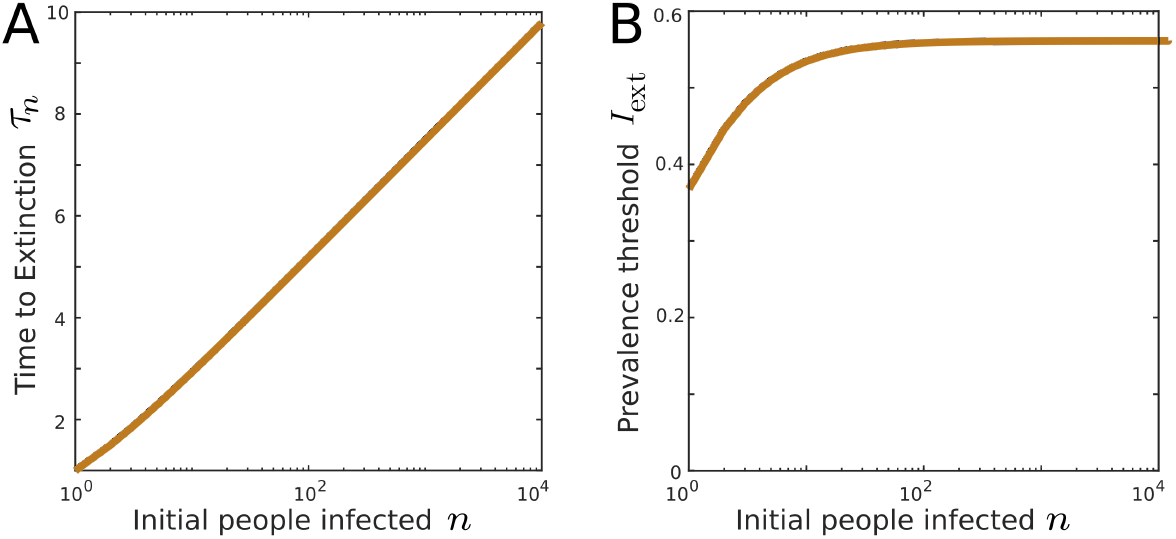
Extinction in a death process model. The average extinction time *τ*_*n*_ and the extinction threshold of infected population *I*_ext_ are plotted as a function of initial number of infected population. Here, *d* = 1 per unit time, with no specific units considered.

Starting with *I*(0) = *n*, the deterministic model predicts an exponential decay of the size of infected population:

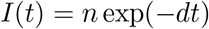

and hence at the expected extinction time we have:

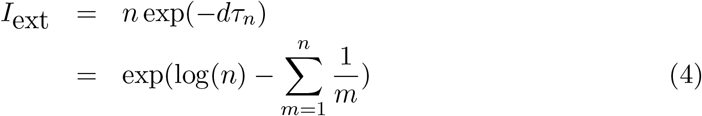

which we postulate gives us a threshold value of the deterministic model that equates to the mean extinction time of the associated stochastic model. Figure 1B shows how the threshold value changes with the initial number of people infected. For large *n*, the threshold plateaus to exp(*−E*), where *E* is Euler’s constant (0.5772) and hence the threshold is 0.5615. Therefore, we observe the useful property that the threshold is largely independent of *n*.

### 2.2 Birth-death dynamics

Moving to the situation where *b* is non-zero, the dynamics are more complex. The expected time to extinction has to be formulated by considering the impact of birth and death from a given number of infected individuals (*n*). This generates an iterated set of equations for the extinction time starting with *n* infections:

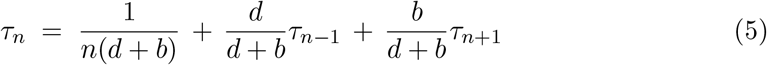

where the first term is the expected time until an event occurs. The other two terms take into account the possibility of a birth or death event transitioning to *n* + 1 or *n −* 1 infected individuals. There is a large relevant literature using similar approaches in epidemiology or in other contexts which lead to comparable equations for the mean extinction time [12–17, 32]. Equation 5 can be expressed as:

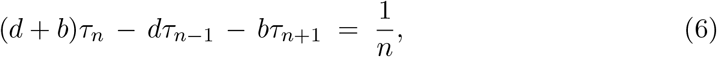

which we solve as a matrix operation (**A***<u>x</u>* = *<u>c</u>*) for *<u>x</u>* = (*τ*_0_ … *τ*_*M*_) with the boundary conditions *τ*_0_ = 0 and 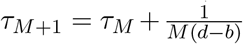 for large *M*, the maximum number of initial people infected.

We note that without loss of generality we can assume that *d* = 1 as this simply acts as a rescaling of time. Figure 2A shows the average extinction time calculated for different choices of *b*. The average extinction time scales with 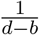 and increases logarithmically with *n* for large values.

**Figure 2:**
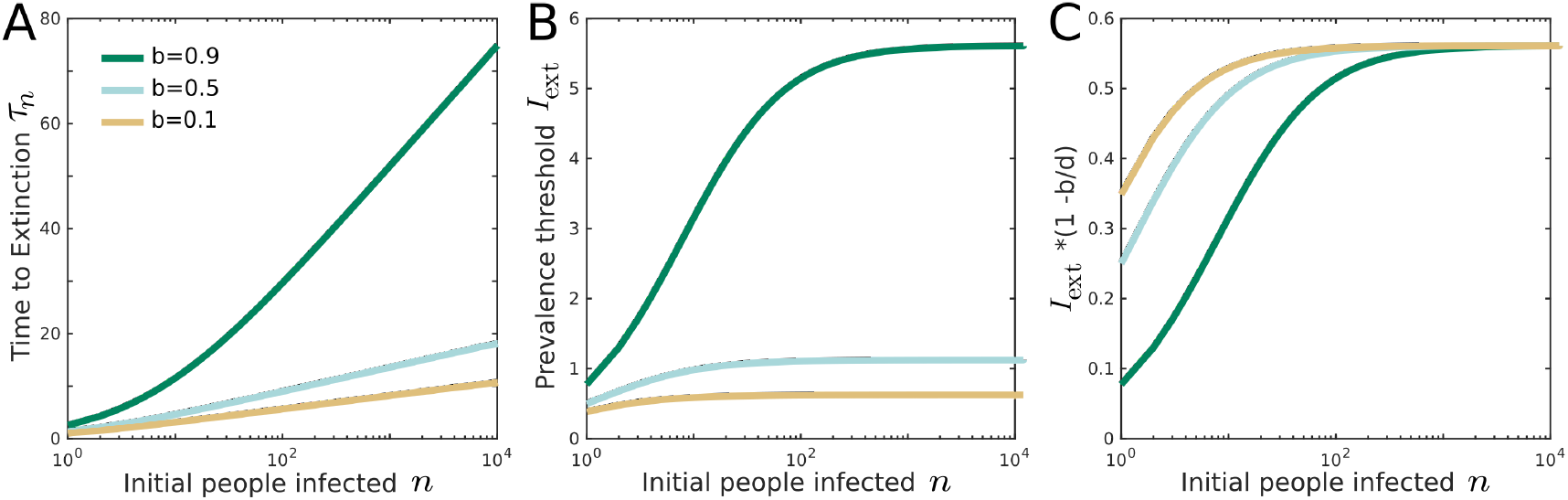
Extinction in a death-birth process model. The average extinction time, *τ*_*n*_, the extinction threshold of infected population, *I*_ext_, and a rescaled version of this threshold are plotted as a function of initial number of infected population for three choices of birth rate *b*. Again, *d* = 1 per unit time.

With the same approach as the previous section, we can calculate the threshold of number of people infected *I*_ext_ = *n* exp((*b − d*)*τ*_*n*_), using the predicted mean extinction time from equation 5. Figure 2B shows how the threshold depends on the initial size of the infected population, calculated for different choices of birth rate *b*. Our results confirm the useful property that the threshold is largely independent of *n* for moderate numbers of initially infected people (e.g. *n >* 500) and that a scaled threshold (by a factor of 1 *− b/d*) reaches the constant value of exp(*−E*) (Figure 2C).

### 2.3 Distribution of extinction times

The above approach to formulate the mean extinction times can be extended to higher cumulative moments to estimate the variation and distribution of extinction time in the birth-death model. The second cumulative moment, 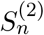, defined as the expected value of the square of the time to extinction, obeys:

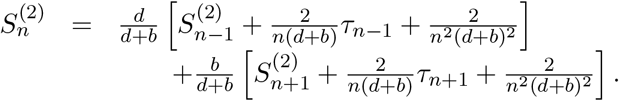

Rearranging the above form, and using our previous expression (Equation 5), we obtain the simplified sequential relation:

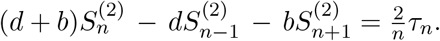

This can be solved with a similar matrix operation as before (**A***<u>x</u>* = *<u>c</u>*^′^) with the boundary conditions *S*_0_ = 0 and 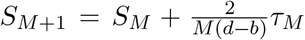 for large *M* to obtain 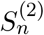 and hence the variance is given by 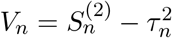

Knowing that the distribution of extinction times is highly asymmetric with a long right-hand tail, higher cumulative moments help provide a more accurate estimate of prediction intervals and the full distribution of extinction times. We find the average cube of extinction times, which is used to calculate the third cumulative moment, obeys:

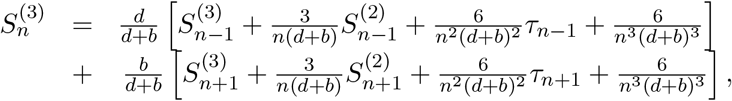

that can be simplified as

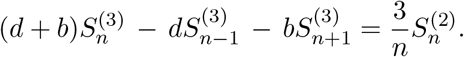

This approach can be extended to calculate sequential relations of cumulative moments of higher orders *m*

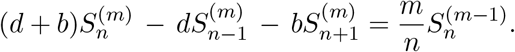

The full set of cumulative moments allows the probability distribution of extinction times to be calculated. Our numerical solutions suggest that the first four moments can give a good estimate of the distribution, approximated by a generalized F probability distribution function with four free parameters. This allows us to determine the prediction intervals for a given set of parameters (details in SI).

As an alternative approach, we consider the Kolmogorov forward equations to describe the time-evolution of probabilities for the numbers infected [33]. These high-dimensional models are a numerically exact realisation of the full stochastic model, which can be computed for SIS model due to the relatively low number of states. The Kolmogorov forward equations are give by:

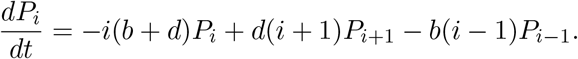

Here *P*_*i*_ depicts the probability of having *i* individuals infected. The solution of this equation is given by **P** = exp (**A***t*) · **P**_**0**_ for the vector of *P*_*i*_s over time, where **A** is the coefficient matrix and **P**_**0**_ is the initial state of the system. We can accordingly calculate the probability of extinction over time and the therefore the distribution.

Figure 3 compares the estimated F distributions of extinction times from the cumulative moments with the corresponding prediction intervals with the solutions of the Kolmogorov forward equations. Again, without loss of generality we have taken *d* = 1, and varied *b < d*. The distribution shifts to the right and flattens as birth rate increases. These results demonstrate the very good agreement between the distributions, as well as the mean values and 95% prediction intervals. This analysis, therefore, provides confidence that our simple approach is a useful tool to estimate extinction times and prediction intervals for any choice of *b* and *d*. In the supplementary information (SI), we plot how variance, skewness, and kortosis change as a function initial population infected *n*.

**Figure 3:**
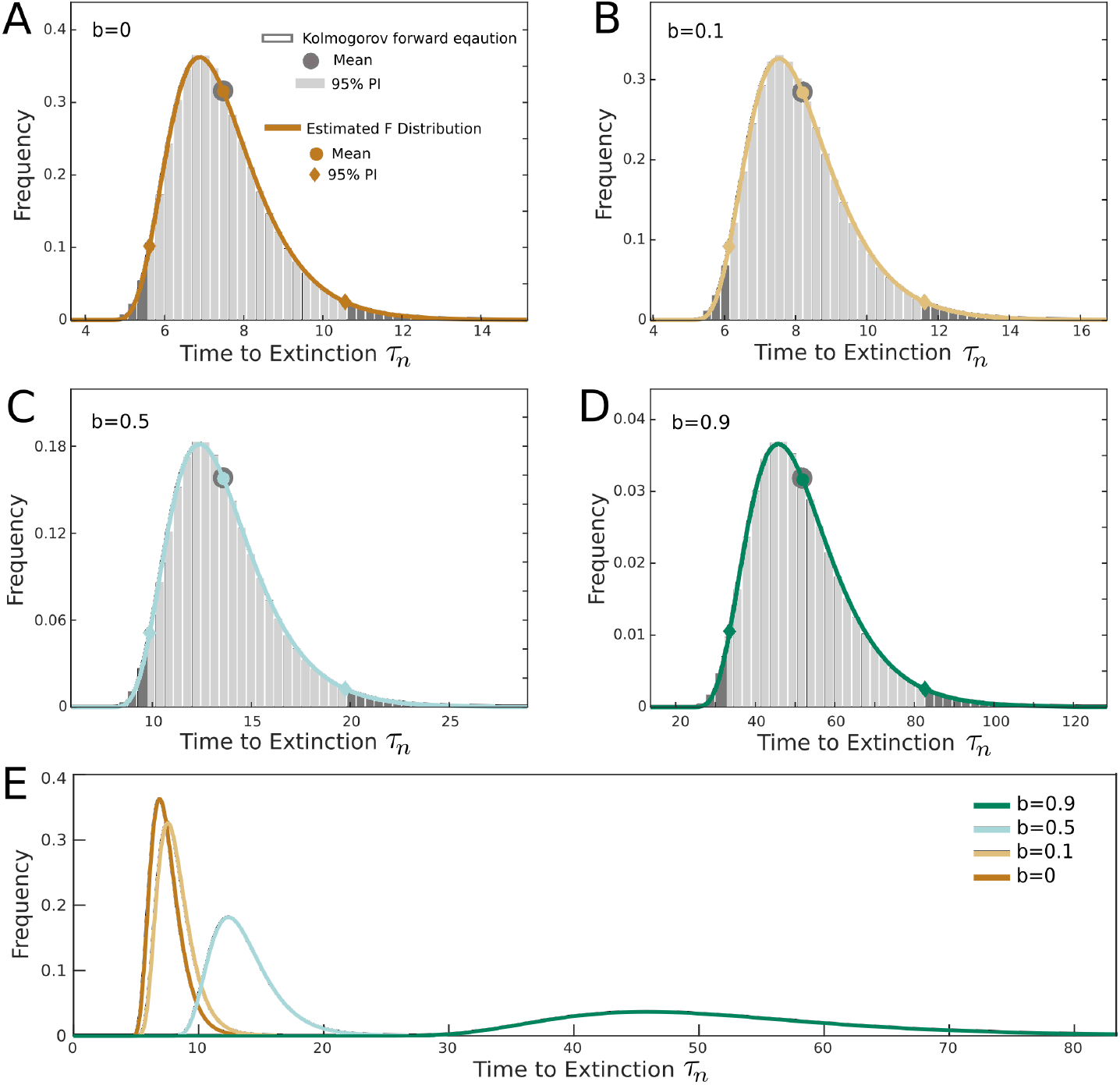
Probability distribution of extinction time *τ*_*n*_ in the model of birth-death process starting with *n* = 1000. (A-D) Gray bars represent the solutions of Kolmogorov forward equation (light gray specifies 95% PI). The colored line shows the F distribution estimated from the first four moments of birth-death process. Corresponding mean values and 95% prediction intervals are indicated for each data set. Each panel corresponds to a birth rate *b*, when *d* = 1 per unit time for all. (E) Comparing the estimated F distribution functions plotted for all values of birth rate *b*.

## 3 Application to the complex dynamics of sleeping sickness

We now investigate how well our theory holds against more realistic, and hence more complex, simulation models. Our previously developed model for gHAT [28, 34] is high dimensional, describing dynamics of vectors and two human risk groups. The model accounts for the natural history of disease progression; infected humans initially develop mild disease (“stage 1”) before progressing to more severe disease (“stage 2”) with neurological symptoms as the parasite crosses the blood-brain barrier. The infectious period ends with either death or successful diagnosis and treatment by screening programs [35]. This model is fitted to the human case incidence data recorded in different administrative regions (health zones of approximately 100,000 people) across the Democratic Republic of Congo to estimate the free parameters of the deterministic model [34].

Using a Markov Chain Monte Carlo approach, posterior parameter sets that match available case data have previously been generated (see [34] and SI for details). The deterministic equations can be numerically solved to generate the disease dynamics including the number of humans infected and new transmission each year for posterior parameter sets. We use these results and our knowledge of the underlying epidemiology to determine appropriate values of *d* and *b* that approximate the linear birth-death process (see SI for details). This allows us to generate the first four moments, and accordingly the distribution of extinction times for specific *d* and *b* rates following the framework developed in the previous sections.

To verify these results, we run the stochastic discrete simulations of the gHAT model using the tau-leaping approach described elsewhere [25, 36] and select a 1-day timestep which approximates the full dynamics well due to the long timescale of gHAT infections (a Gillespie method could be used for more accurate simulations, however it is unlikely to modify the results we present here). We compute the extinction times for multiple stochastic realisations. These simulations assume that active screening (the fundamental driver of case identification and control) continues at a constant level, leading to decaying infection levels (Figures in the SI). Figure 4 compares the F probability distribution function derived from the estimated cumulants with the distribution of extinction times of one million stochastic simulations for two different parameter sets. The parameter sets represent the most likely of all posterior parameter sets found by fitting to data for two health zones named Mosango (a “moderate-risk” zone) and Kwamouth (a “high-risk” zone) [34] that reveal different infection dynamics over time, and therefore different predicted extinction times. Despite the complexity of the model for gHAT dynamics, there is still a very good agreement between the birth-death model with F distribution predictions and full stochastic gHAT infection dynamics (with differences in mean and 95% PI of less than a year) confirming the analysis based on birth-death model can deliver robust and accurate estimates of mean extinction time and the prediction intervals. Examples using other levels of screening coverage are shown in the SI. In these examples, for a fixed parameter set, we computed that the deterministic threshold, *I*_ext_, to compute the expected extinction time was 1.39 and 1.22 for Mosango and Kwamouth respectively. Other threshold values would need to be calculated for alternative parameter sets and alternative screening strategies for the same health zones.

It should be noted that this study provides a simplified picture of eradication of a diseases taking into account instant feedback between transmission and the infected population. However in epidemiology, other measures such as elimination of transmission may be of interest. In the case of complex dynamics like gHAT, due to phase difference of humans and vectors as well as slow progression of the disease and recovering process, there is a delay between elimination of transmission and disease extinction. We speculate the lag period can be estimated based on the underlying dynamics.

**Figure 4:**
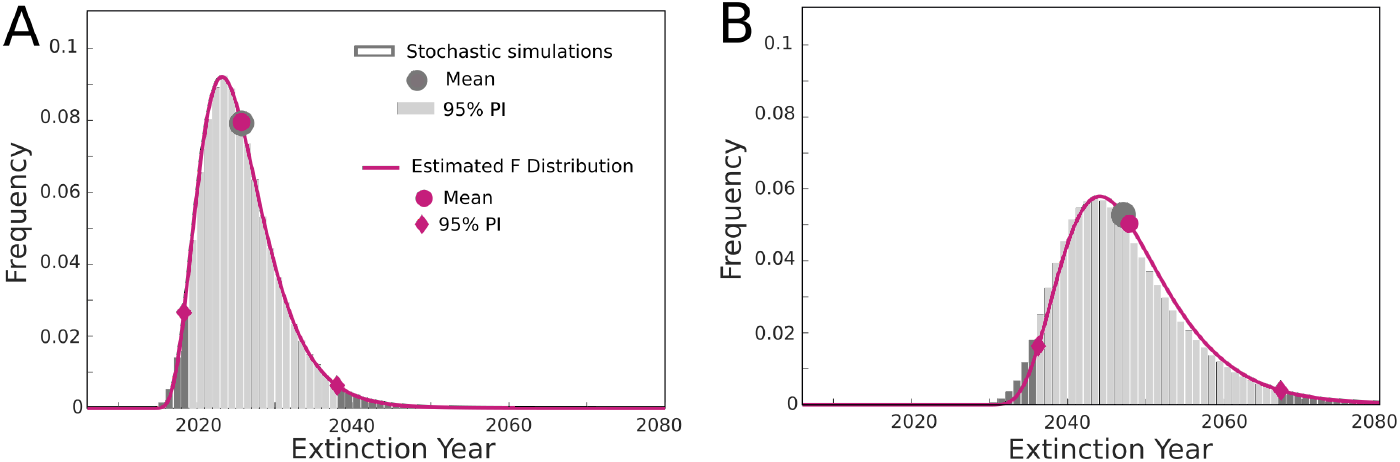
Probability distribution of extinction time for the full (17-dimensional) gHAT model for two health zones, (A) Mosango and (B) Kwamouth with 30% active screening. Gray bars represent the solutions of one million stochastic simulations (light gray specifies 95% PI). The purple line shows the F distribution estimated from the first four moments of the simplified birth-death process approach (*b* = 0.38, *d* = 0.64 for Mosango and *b* = 0.25, *d* = 0.46 for Kwamouth). Corresponding mean values and 95% prediction intervals are indicated for each data set.

Another challenge is to extend such analysis to allow for more general noise models, while our current stochastic models mainly account for Poisson events. In the case of gHAT dynamics, we are aware of possible over-dispersion terms that can be considered in the infection dynamics [25,34]. It is important to study how the predicted distribution of extinction time will be modified by such noise models.

## 4 Conclusion

In this manuscript, we study the extinction times of diseases when *R*_eff_ *<* 1 starting from a finite population infected, utilising results from a simplified birth-death model. We formulate the mean extinction time that is in agreement with the previous studies in the analogous limits [13–17]. Beyond that, our analysis provides a novel method to approximate the distribution of extinction times by calculating additional moments of extinction time. Our results agree well with the solutions of forward Kolmogorov equations, which offer a numerical exact prediction of the birth-death dynamics. Our findings are a vital extension to deterministic models, allowing a robust and rigorous assessment of extinction in a modelling framework that cannot naturally capture this phenomena. We considered the simplistic assumption of constant birth and death rates, however, the results can be easily extended by modifying the coefficient matrix when these rates are functions of infected population.

Moreover, our analysis helps us to quantify a simple threshold that can be used to determine the mean extinction time from deterministic solutions. We showed that this threshold asymptotes to a universal plateau given by effective reproduction ratio for large initial numbers of infected individuals. This asymptotic threshold is likely to be a reasonable approximation for large populations where the number of infections is also likely to be large.

Interestingly, the birth-death approach can be applied to diseases with more complex disease dynamics in order to predict their extinction times. We showed that it is valid for the example of local elimination of sleeping sickness, yet in a broad sense the dynamics are approximately SIS decaying to zero infection. Our study can be extended for SIR models, where the system state is two-dimensional by considering the recovered compartment. A non-trivial modification of the birth-death process would be helpful to consider to understand the richer dynamics of the SIR model [37].

## Data Availability

All the necessary data and details of methods are included in the supplementary information.

## Acknowledgements

The authors thank Dr Erick Mwamba Miaka and PNLTHA for original data collection, and WHO for data access (in the framework of the WHO HAT Atlas [27]). This work was supported by the Bill and Melinda Gates Foundation (www.gatesfoundation.org) through the NTD Modelling Consortium [OPP1184344] (M.A., K.S.R. and M.J.K.) and the Human African Trypanosomiasis Modelling and Economic Predictions for Policy (HAT MEPP) project [OPP1177824] (K.S.R. and M.K.). The funders had no role in study design, data collection and analysis, decision to publish, or preparation of the manuscript.

## Author contribution

All authors contributed in developing the model and writing the manuscript.

## Data Accessibility

This work does not include any original data. To describe the gHAT dynamics, we used the model and the fitted parameters from a published manuscript which is cited in the main text and discussed in details in the supplementary material.

