## Supplementary information for "Estimating the distribution of time to extinction of infectious diseases in mean-field approaches"

Maryam Aliee, Kat S. Rock, Matt J. Keeling
Mathematics Institute, University of Warwick, Coventry, CV4 7AL, UK
Zeeman Institute for Systems Biology and Infectious Disease Epidemiology Research,
University of Warwick, Coventry, CV4 7AL, UK

Content:

- Supplementary figures and tables
  - Figure 1 - Moments of extinction time in a death-birth process model.
  - Figure 2 - Probability distribution of extinction time for the full gHAT model.
  - Figure 3 - Solutions of the deterministic model for gHAT dynamics.
  - Table 1 - Estimated birth-death model parameters of the gHAT model.
- Methods
  - Estimating the distribution of extinction time
  - Description of the gHAT model
- References

### 1 Supplementary figures

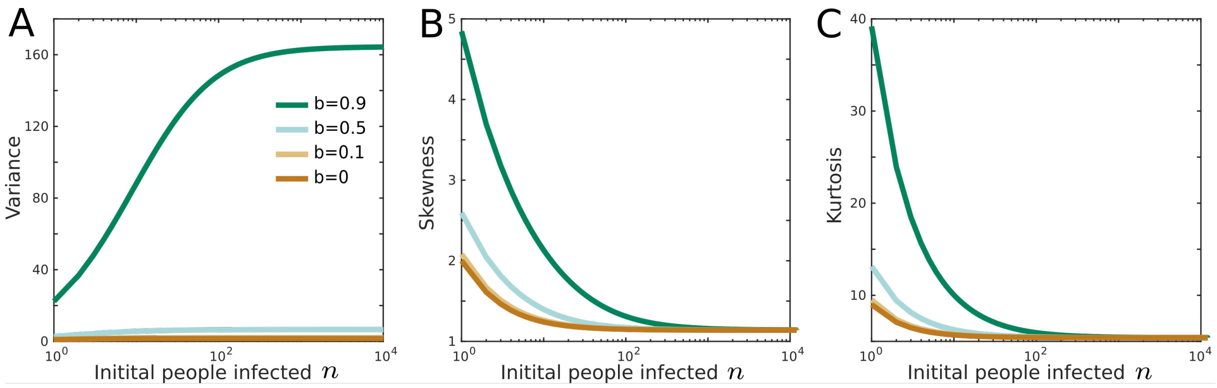


Figure 1: Moments of extinction time in a death-birth process model. Variance, skewness and kurtosis of extinction time are plotted as a function of initial number of infected population for different choices of birth rate $b$, $d = 1$.

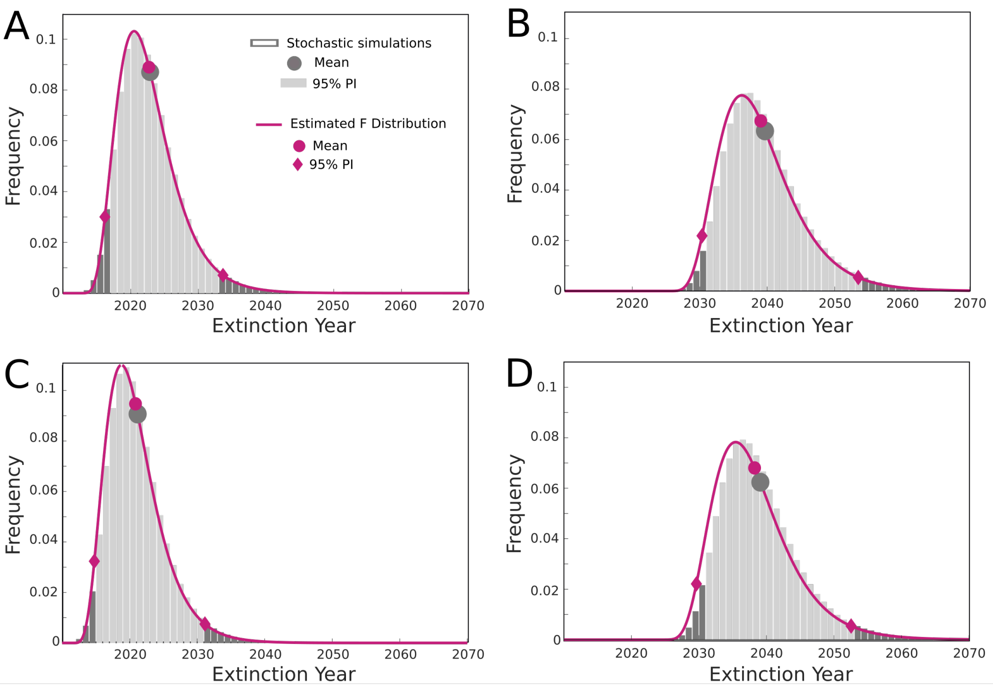


Figure 2: Probability distribution of extinction time for the full gHAT model for two health zones, (A,C) Mosango and (B,D) Kwamouth. First and second lines respectively correspond to 40% and 50% active screening. Gray bars represent the solutions of one million stochastic simulations (light gray specifies 95% PI). The purple line shows the F distribution estimated from the first four moments of birth-death process. Corresponding mean values and 95% prediction intervals are plotted for each data set.

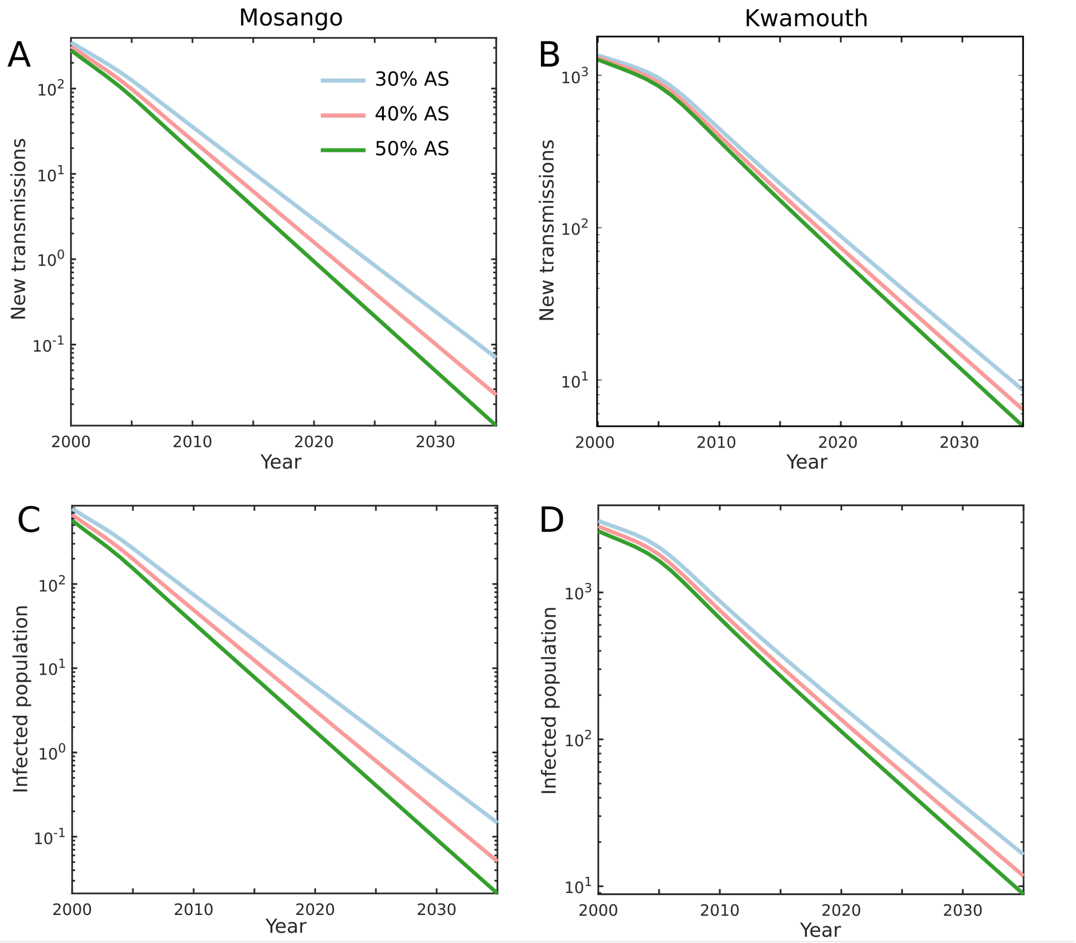


Figure 3: Solutions of the deterministic model for gHAT dynamics. Annual number of new transmissions and the people infected are plotted over time for two health zones. Different colours represent three levels of active screening used in the model.

Table 1: Estimated birth-death model parameters of the gHAT model for two health zones Mosango and Kwamouth and with different screening levels.

| **Active screening** | **Mosango** | **Kwamouth** |
|---|---|---|
| 30% | $b = 0.42$<br>$d = 0.67$<br>$I_{\text{ext}} = 1.50$ | $b = 0.48$<br>$d = 0.64$<br>$I_{\text{ext}} = 2.60$ |
| 40% | $b = 0.43,$<br>$d = 0.71$<br>$I_{\text{ext}} = 1.45$ | $b = 0.50$<br>$d = 0.66$<br>$I_{\text{ext}} = 2.24$ |
| 50% | $b = 0.45$<br>$d = 0.75$<br>$I_{\text{ext}} = 1.41$ | $b = 0.51$<br>$d = 0.69$<br>$I_{\text{ext}} = 2.23$ |

### 2 Estimating the distribution of extinction time

The probability distribution function of a random variable can be identified in a unique way with the full set of cumulative moments [1]. For practical reasons, numerical methods can be used to estimate the distribution function with the help of the first few moments. In our analysis of the birth-death process, MATLAB fitting functions suggest the modified F probability distribution can capture the distribution of extinction time calculated in our model. Generalized F distribution is a shifted scaled version of F distribution defined as

$$PDF(t) = F(\frac{t+C}{M};\ \nu_1, \nu_2),$$

which has four free parameters. Therefore, we use the first four moments calculated in section 2.2 in the main text to estimate the four parameters of the F distribution. Mean $\tau_n$, variance $S_n^2 - \tau_n^2$, skewness $S_n^3 - 3\tau_n S_n^2 + 2\tau_n^3$, and kurosis $S_n^4 - 4\tau_n S_n^3 + 6\tau_n^2 S_n^2 - 3\tau_n^4$ are matched to the corresponding values of the probability distribution function to estimate the four parameters of $C$, $M$, $\nu_1$, and $\nu_2$. The mean and 95% prediction intervals are then calculated from the cumulative distribution straightaway.

### 3 Description of the gHAT model

Our model to describe gHAT dynamics is based on the ODE model presented in [2, 3] and the extended stochastic version in [4, 5]. Figure 4 shows a schematic description of gHAT dynamics in this model that takes into account different compartments of humans and tsetse. Humans can be exposed and subsequently infected by a bite of an infectious tsetse. They progress through stage 1 and stage 2 of the infection with specific rates. On the other side, tsetse vectors can become exposed and subsequently infectious if they bite an infectious human. Infected people may be detected by passive and active screening (see below for more details), followed by hospitalisation and recovery. Here, we consider a version of the model where humans are partitioned into two sub-groups of (i) low-risk and participating in the active screening, and (ii) high-risk and non-participating in active screening. We assume there are no animal reservoirs although animals receive some proportion of tsetse bites. For simplicity, we assume the total population of humans to be constant.

This model accounts for the possibility of detecting of infected humans through passive surveillance and active screening. In this work, we assume a constant ratio (between 30-50%) of active screening from 1998 on, although in the original model the screening level is fixed by the available data.

Passive surveillance describes potential visits of people to fixed medical centers for testing. This is considered in the model with the rates proportional to $\eta_H(Y)$ and $\gamma_H(Y)$ corresponding to the first and second stages of the disease. Before 1998 (pre-active screening) it was assumed that passive detection was less effective than after activities began, and only so identified stage 2 individuals at a rate $\gamma_H^{\text{pre}}$, which is smaller than the stage 2 passive detection rate in 1998, $\gamma_H^{\text{post}}$. Following previous modelling work using gHAT data from former Bandundu province, there is a strong signal from epidemiological staging data that passive screening has improved during the time period from 2000–2012 [2, 6]. To capture the improvement of stage 1 to stage 2 passive detection, the model utilises the following formula:

$$\eta_H(Y) = \eta_H^{\text{post}} \left[ 1 + \frac{\eta_H \text{amp}}{1 + \exp\left(-d_{\text{steep}}(Y - d_{\text{change}})\right)} \right],$$

$$\gamma_H(Y) = \gamma_H^{\text{post}} \left[ 1 + \frac{\gamma_H \text{amp}}{1 + \exp\left(-d_{\text{steep}}(Y - d_{\text{change}})\right)} \right],$$

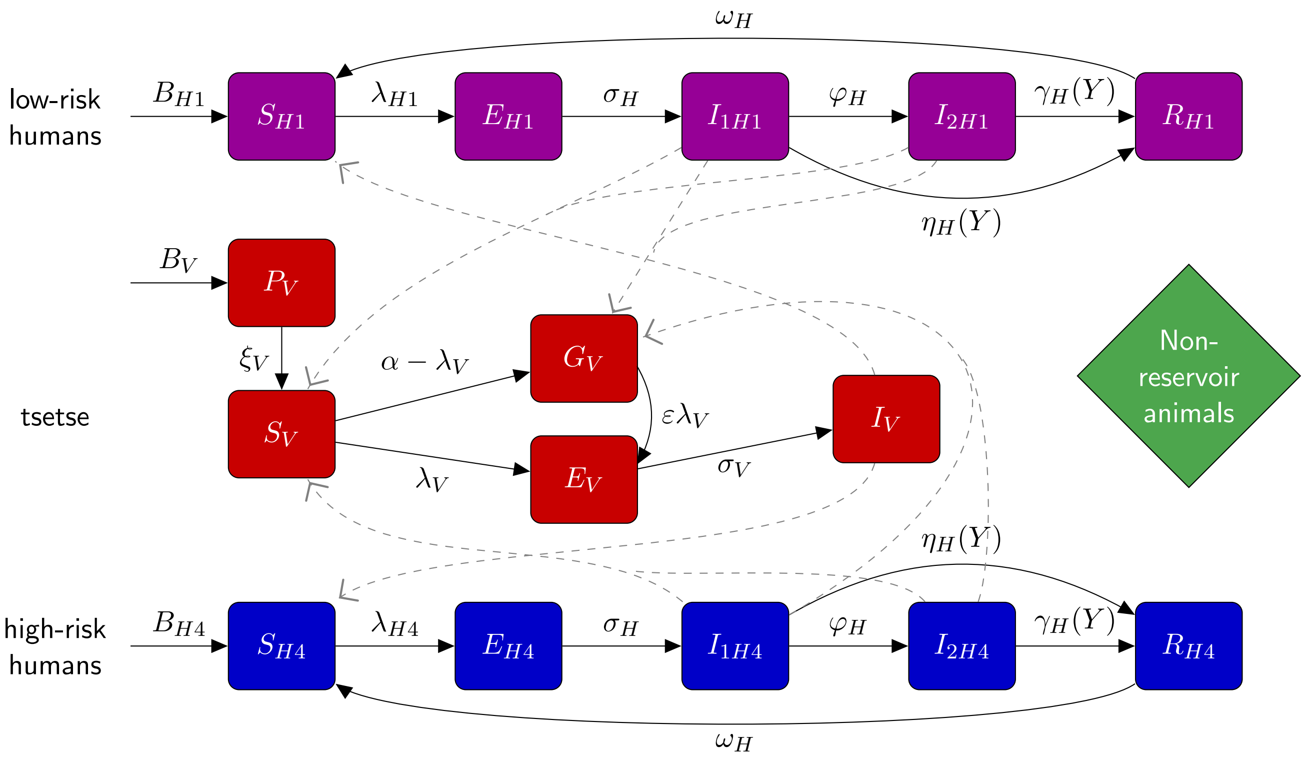


Figure 4: **Schematic of the model to describe gHAT infection dynamics.** This multi-host model of HAT takes into account high- and low-risk groups of humans and their interactions with tsetse vectors. Each group consists of different compartments: Susceptible humans $S_{Hi}$ can become exposed on a bite of an infectious tsetse. Exposed people $E_{Hi}$ progress to become the stage $1$ infected people and eventually stage $2$ (if not detected in screening), and once treated they recover by hospitalization $R_{Hi}$. Active screening can accelerate treatment rate of infected people. Here we assume high-risk group does not participate in active screening. By biting an infectious person, tsetse can become exposed and subsequently infectious, $E_V$ and $I_V$. $G_V$ represents the tsetse population not exposed to *Trypanosoma brucei gambiense* in the first blood-meal and are therefore less susceptible in the following meals. Rates are shown by Greek letters associated with arrows. Animal reservoir is not considered. This figure is taken from [2] and adapted from the original model schematic [3].

where $Y$ is the year and $\eta_H(Y)$ is the annual stage 1 passive detection rate. Parameters dictating the amplitude, steepness and switching year can be found in Tables 2 and 3. The free parameters have been estimated through fitting to the health zone level data for Mosango and Kwamouth.

#### 3.1 Deterministic model

In the original deterministic framework, infection dynamics is described by a set of ODEs. Dynamics of human compartments are given by

$$\begin{aligned}
\frac{dS_{Hi}}{dt} &= \mu_H N_{Hi} - \mu_H S_{Hi} + \omega_H R_{Hi} + \omega_{IA} I_{1Ai} - \alpha m_{\text{eff}} f(i) \frac{S_{Hi}}{N_{Hi}} I_V \\
\frac{dE_{Hi}}{dt} &= \alpha m_{\text{eff}} f(i) \frac{S_{Hi}}{N_{Hi}} I_V - (\sigma_H + \mu_H) E_{Hi}, \\
\frac{dI_{1Hi}}{dt} &= \sigma_H E_{Hi} - (\varphi_H + \mu_H + \eta_H(Y)) I_{1Hi}, \\
\frac{dI_{2Hi}}{dt} &= \varphi_H I_{1Hi} - (\gamma_H(Y) + \mu_H) I_{2Hi}, \\
\frac{dR_{Hi}}{dt} &= \eta_H(Y) I_{1Hi} + \gamma_H(Y) I_{2Hi} - (\omega_H + \mu_H) R_{Hi}, \qquad (3.1)
\end{aligned}$$

where $i$ refers to high or low risk population. The set of equations for tsetse is given by Equations 3.2.

$$\begin{aligned}
\frac{dS_V}{dt} &= \mu_V N_H - \alpha S_V - \mu_V S_V \\
\frac{dE_{1V}}{dt} &= \alpha p_V \Big(f_{H1} \frac{I_{1H1} + I_{2H1}}{N_{H1}} + f_{H2} \frac{I_{1H2} + I_{2H2}}{N_{H2}}\Big)(S_V + \varepsilon G_V) - (3\sigma_V + \mu_V) E_{1V} \\
\frac{dE_{2V}}{dt} &= 3\sigma_V E_{1V} - (3\sigma_V + \mu_V) E_{2V} \\
\frac{dE_{3V}}{dt} &= 3\sigma_V E_{2V} - (3\sigma_V + \mu_V) E_{3V} \\
\frac{dI_V}{dt} &= 3\sigma_V E_{3V} - \mu_V I_V \\
\frac{dG_V}{dt} &= \alpha \left(1 - p_V \left(f_{H1} \frac{I_{1H1} + I_{2H1}}{N_{H1}} + f_{H2} \frac{I_{1H2} + I_{2H2}}{N_{H2}}\right)\right) S_V \\
&- \alpha p_v \left(f_{H1} \frac{I_{1H1} + I_{2H1}}{N_{H1}} + f_{H2} \frac{I_{1H2} + I_{2H2}}{N_{H2}}\right) \varepsilon G_V - \mu_V G_V. \qquad (3.2)
\end{aligned}$$

These deterministic equations are numerically solved with the help of Runge-Kutta methods to find the disease dynamics including the number of humans infected and new transmission each year (Figure3). We use these results to impute appropriate values of $d$ and $b$ that approximate the linear birth-death process. Infected population or number of new transmission would decrease since $R_{eff} < 1$ with an effective rate given by $d - b$ in time in the logarithmic scale. We approximate $d$ as the number of new transmission per infected population of humans.

#### 3.2 Estimating model parameters

As in previous versions of the deterministic model [2, 3], some parameters with estimates available in the literature were assigned fixed values. Fixed values are given in Table 2. The other parameter values were taken from posterior distributions by fitting the model to data. In this approach, the deterministic model was fitted to health-zone-level data for different health zones using an adaptive Metropolis-Hastings MCMC algorithm [2]. In the present analysis, we choose the most likely parameter sets from the posterior distributions for two different health zones of Mosango and Kwamouth.

Table 2: **Model parameterisation (fixed parameters).** Notation, a brief description, and the used values of fixed parameters in model W.

| Notation | Description | Value |
|---|---|---|
| $N_H$ | Total human population size (in 2015) | M:121,433 K1:127,205 [7] |
| $B_H$ | Total human birth rate | $= \mu_H N_H$ |
| $\mu_H$ | Natural human mortality rate | 5.4795$\times 10^{-5}$ days$^{-1}$ [8] |
| $\sigma_H$ | Human incubation rate | 0.0833 days$^{-1}$ [9] |
| $\varphi_H$ | Stage 1 to 2 progression rate | 0.0019 days$^{-1}$ [10, 11] |
| $\omega_H$ | Recovery rate/waning-immunity rate | 0.006 days$^{-1}$ [12] |
| Sens | Active screening diagnostic sensitivity | 0.91 [13] |
| $B_V$ | Tsetse birth rate | 0.0505* days$^{-1}$ [14] |
| $\xi_V$ | Pupal death rate | 0.037 days $^{-1}$ |
| $K$ | Pupal carrying capacity | $= 111.09 N_H^{\dagger}$ [14] |
| $\mathbb{P}$(pupating) | Probability of pupating | 0.75 |
| $\mu_V$ | Tsetse mortality rate | 0.03 days$^{-1}$ [9] |
| $\sigma_V$ | Tsetse incubation rate | 0.034 days$^{-1}$ [15, 16] |
| $\alpha$ | Tsetse bite rate | 0.333 days$^{-1}$ [17] |
| $p_V$ | Probability of tsetse infection per single infective bite | 0.065 [9] |
| $\varepsilon$ | Reduced non-teneral susceptibility factor | 0.05 [3] |
| $f_H$ | Proportion of blood-meals on humans | 0.09 [18] |

#### 3.3 Stochastic model

We also use the stochastic version of the described gHAT model to study the extinction time [4, 5]. In this stochastic picture, individual humans are assigned to different compartments associated with infection/disease status and can transfer between them. We describe system dynamics by random events captured by a tau-leap approximation. Table 4 explains different events and the corresponding rates that lead to one person transitioning from one compartment to another one. Within this framework, the number of events happening in a time interval $\tau$ is chosen randomly from a Poisson distribution with the mean equal to the event rate multiplied by $\tau$. The same procedure is used to identify the number of people participating in passive or active screenings. We use time interval of a day in the tau-leaping algorithm that is shown to be a sensible choice for gHAT dynamics [4].

For simplicity reasons, we keep vector dynamics the same as the original model, described by a set of ODEs. This is a legitimate assumption due to the high population of vectors and their short life cycle compared to humans.

For each, we perform 1 million realisations to achieve reasonable statistics of the extinction time.

Table 3: **Model parameterisation (posterior parameters).** Notation, a brief description, and the values used for two health zones Mosango and Kwamouth.

| Notation | Description | Value | | Unit |
|---|---|---|---|---|
| | | Mosango | Kwamouth | |
| $R_0$ | Basic reproduction number (NGM approach) | 1.015 | 1.07 | - |
| $r$ | Relative bites taken on high-risk humans | 3.322 | 6.213 | - |
| $k_1$ | Proportion of low-risk people | 0.909 | 0.888 | - |
| $k_4$ | Proportion of high-risk people | $k_4 = 1 - k_1$ | | - |
| $\eta_H^{\text{post}}$ | Treatment rate from stage 1 (1998) | 8.646$\times 10^{-5}$ | 1.147$\times 10^{-4}$ | days$^{-1}$ |
| $\gamma_H^{\text{pre}}$ | Treatment rate from stage 2 (pre-1998) | 1.906 $\times 10^{-3}$ | 1.947$\times 10^{-3}$ | days$^{-1}$ |
| $\gamma_H^{\text{post}}$ | Treatment rate from stage 2 (1998) | 2.574$\times 10^{-3}$ | 1.961$\times 10^{-3}$ | days$^{-1}$ |
| Spec | Active screening diagnostic specificity | 0.9994 | 0.9992 | - |
| $u$ | Proportion of passive cases reported | 0.2756 | 0.2477 | - |
| $\eta_{H\text{amp}}$ | Relative improvement in passive stage 1 detection rate | 0.752 | 2.099 | - |
| $\gamma_{H\text{amp}}$ | Relative improvement in passive stage 2 detection rate | 0.287 | 0.536 | - |
| $d_{\text{steep}}$ | Speed of improvement in passive detection rate | 1.190 | 0.857 | - |
| $d_{\text{change}}$ | Midpoint year for passive improvement | 2004.3 | 2005.5 | - |


[3] Kat S Rock, Steve J Torr, Crispin Lumbala, and Matt J Keeling. Quantitative evaluation of the strategy to eliminate human African trypanosomiasis in the DRC. *Parasites & Vectors*, 8(1):532, 2015.

[4] Christopher N Davis, Kat S Rock, Erick Mwamba Miaka, and Matt J Keeling. Village-scale persistence and elimination of gambiense human African trypanosomiasis. *medRxiv*, page 19006502, 2019.

[5] M Soledad Castaño, Maryam Aliee, Erick Mwamba Miaka, Matt J Keeling, Nakul Chitnis, and Kat S Rock. Screening strategies for a sustainable endpoint for gambiense sleeping sickness. *The Journal of Infectious Diseases*, page jiz588, 2019.

[6] M Soledad Castaño, Martial L Ndeffo-Mbah, Kat S Rock, Cody Palmer, Edward Knock, Erick Mwamba Miaka, Joseph M Ndung'u, Steve Torr, Paul Verlé, Simon E F Spencer, and Others. Assessing the impact of data aggregation in model predictions of HAT transmission and control activities. *medRxiv*, page 19005991, 2019.

[7] OCHA Office for the Coordination of Humanitarian Affairs. *Journées Nationales de Vaccination (JNV) Activités de vaccination supplémentaire , RDC*, Accessed May 2016.

[8] World Development Indicators. World bank. Technical report, 2015.

[9] DJ Rogers. A general model for the African trypanosomiases. *Parasitology*, 97(1):193–212, 1988.

[10] Francesco Checchi, João A N Filipe, Daniel T Haydon, Daniel Chandramohan, and François Chappuis. Estimates of the duration of the early and late stage of gambiense sleeping sickness. *BMC infectious diseases*, 8(1):16, 2008.

Table 4: Model formulation (human dynamics)

| **Event** | **Transition** | **Rate** |
|---|---|---|
| Recovery from hospitalisation | $s_{Hi} \rightarrow s_{Hi} + 1, r_{Hi} \rightarrow r_{Hi} - 1$ | $\omega_H r_{Hi}$ |
| Natural death of hospitalised | $s_{Hi} \rightarrow s_{Hi} + 1, r_{Hi} \rightarrow r_{Hi} - 1$ | $\mu_H r_{Hi}$ |
| Exposure of susceptibles | $s_{Hi} \rightarrow s_{Hi} - 1, e_{Hi} \rightarrow e_{Hi} + 1$ | $f_{Hi} \alpha m_{eff} s_{Hi} I_V / N_{Hi}$ |
| Progression to Stage 1 infection | $e_{Hi} \rightarrow e_{Hi} - 1, i_{1Hi} \rightarrow i_{1Hi} + 1$ | $\sigma_H e_{Hi}$ |
| Natural death of exposed | $e_{Hi} \rightarrow e_{Hi} - 1, s_{Hi} \rightarrow s_{Hi} + 1$ | $\mu_H e_{Hi}$ |
| Progression to Stage 2 infection | $i_{1Hi} \rightarrow i_{1Hi} - 1, i_{2Hi} \rightarrow i_{2Hi} + 1$ | $\varphi_H i_{1Hi}$ |
| Natural death of Stage 1 infection | $i_{1Hi} \rightarrow i_{1Hi} - 1, s_{Hi} \rightarrow s_{Hi} + 1$ | $\mu_H i_{1Hi}$ |
| Treatment or death from Stage 2 infection | $s_{Hi} \rightarrow s_{Hi} + 1, i_{2Hi} \rightarrow i_{1Hi} - 1$ | $\gamma_H(Y) i_{2Hi}$ |
| Natural death of Stage 2 infection | $i_{2Hi} \rightarrow i_{2Hi} - 1, s_{Hi} \rightarrow s_{Hi} + 1$ | $\mu_H i_{2Hi}$ |

[17] World Health Organization. Control and surveillance of human african trypanosomiasis: report of a who expert committee. Technical report, 2013.

[18] PH Clausen, I Adeyemi, B Bauer, M Breloeer, F Salchow, and C Staak. Host preferences of tsetse (Diptera: Glossinidae) based on bloodmeal identifications. *Medical and Veterinary Entomology*, 12(2):168–180, 1998.
